# System pressures may threaten patient perceptions and experiences of empathy in primary care consultations: A nested qualitative interview study

**DOI:** 10.64898/2026.08.24.26361185

**Authors:** Rachel Dewar-Haggart, Emma Teasdale, Sebastien Pollet, Geraldine M Leydon, Hazel A Everitt, Leanne Morrison, Helen Atherton, Jeremy Howick, Isla Davis, Shola Falohun, Jennifer Bostock, Jane Vennik, Nadia Cross, Paul Little, Christian D Mallen, Matthew J Ridd, Amy Herbert, Michelle E Robinson, Jacqueline Nuttall, Taeko Becque, Kirsty Garfield, Beth Stuart, Nazrul Islam, Paul H Lee, Felicity L Bishop

**Author notes:** Correspondence to: R Dewar-Haggart. Nuffield Department of Primary Care Health Sciences, University of Oxford, Radcliffe Observatory Quarter, Woodstock Road, Oxford, OX2 6GG.

## Abstract

**Background:** Effective communication during consultations is facilitated by clinical empathy and realistic optimism, and can enhance patient satisfaction with care, alleviate symptoms, and improve quality of life. However, primary care systems are under significant strain and changing rapidly, which may affect practitioners’ ability to communicate empathically and convey realistic optimism, with implications for the patient-practitioner relationship and patient outcomes. Understanding patients’ perspectives of healthcare communication in the current clinical context is therefore important. We aimed to explore patients’ experiences and perceptions of communication in UK primary care consultations, focussing on the communication of clinical empathy and realistic optimism.

**Methods:** A qualitative interview study was conducted as part of a multi-centre cluster-randomised trial of EMPathicO, a brief e-learning package for Primary Care Practitioners (PCPs) on communicating clinical empathy and realistic optimism. Participants were not aware whether their general practice had access to EMPathicO or not. Interviews were conducted within 7-14 days of participants’ consultations, explored their views and experiences of clinical empathy and realistic optimism, and were transcribed verbatim. Interviews were analysed using Ritchie and Spencer’s Framework Method.

**Results:** We conducted semi-structured audio-recorded qualitative telephone interviews with 71 participants from 29 primary care practices taking part in the EMPathicO trial. Following comprehensive mapping of data to the framework derived following initial analysis, four themes were agreed. Overall, most participants described positive empathic consultations with their PCPs, however, participants’ experiences were shaped by wider systemic and contextual factors. They described a stretched and inefficient primary care system impacting empathy and optimism; the impact of PCP ‘preparedness’ as a marker for empathy; how consultation modality (i.e. in-person or telephone) shaped perceptions of empathy, and how PCPs sharing next steps in participants’ treatment and management could foster realistic optimism.

**Conclusions:** While clinical empathy and realistic optimism may be experienced by patients during consultations with practitioners, the wider contextual challenges of accessing and navigating primary care systems can threaten overall perceptions of feeling cared for. Future primary care policy and workforce training must consider these system pressures to preserve effective communication in consultations and positive patient-practitioner encounters.

**Trial registration:** ISRCTN18010240 registered 15^th^ September 2022.

## Background

Effective communication between primary care practitioners (PCPs) and patients can enhance patient satisfaction with care, alleviate symptoms, and improve overall quality of life.(1–3) This communication can be facilitated through clinical empathy, which involves the PCP considering the patient’s viewpoint by exploring and acknowledging their feelings, concerns, and expectations, while maintaining professional boundaries.(4, 5) Moreover, communicating realistic optimism, which involves communicating confidence in a suggested treatment or management plan (such as prescribed medication or lifestyle changes) may lead to improvement in function or other desired patient outcomes, and is considered to be part of clinical empathy.(6) Using clinical empathy can foster a patient’s sense of trust in their PCP, and conveying positive messages, as long as they are realistic and personalised to the patient, can enhance consultations for osteoarthritis and other chronic conditions.(7) However, primary care in the UK and worldwide is under significant strain and systems are changing rapidly(8), which may affect the delivery of empathic care. There has been a wholescale change in how consultations are conducted in primary care, with a move to more use of remote methods and access to a wider range of healthcare professionals beyond GPs, including First Contact Practitioners (FCPs); healthcare professionals who provide the first point of contact and assessment for defined groups of patients and presentations.(9–12) However, the impact of these changes on patients’ experiences have been overlooked. Patients want relationships with their chosen doctor rather than just fast access to care, and this may be compromised by these system-wide changes.(13) Therefore, understanding patient perspectives of healthcare communication in the current primary care landscape is important.(13)

Talking in Primary Care (TIP) was a cluster-randomised controlled trial to assess clinical and cost-effectiveness of an eLearning tool (EMPathicO), which aimed to enhance Primary Care Practitioners’ (PCP) communication of clinical empathy and realistic optimism in primary care consultations.(14) This nested qualitative study with patient participants enabled an exploration of patients’ experiences and perceptions of communication in primary care consultations.

## Methods

### Aims & Objectives

The aim of this nested qualitative study was to explore patients’ experiences and perceptions of primary care consultations, focussing on communication of clinical empathy and realistic optimism.

### Design

We approached the mixed methods project with a pragmatist epistemology, seeking to generate knowledge that can make a difference to practice by selecting appropriate methods for specific aims and applying them with rigour.(15) To explore patients’ experiences and perceptions we conducted one-to-one semi-structured telephone interviews with a diverse sample of participants and applied framework analysis.(16, 17) Telephone interviews were chosen to reduce participant burden and enable wider geographical reach within the context of this nationwide trial. The Framework Method was chosen to facilitate the process of sifting, charting, and sorting large amounts of interview data into key themes with potential policy implications. The qualitative team included two female qualitative research fellows (RDH, ET), a female health psychologist (FLB), a female psychologist (LM), a male research fellow (SP), a female medical sociologist (GL) and a female general practitioner (HAE).

### Setting, recruitment, and participants

Participants were recruited from 53 primary care practices across England and Wales who took part in a multi-centre cluster-randomised controlled trial of EMPathicO, an e-learning package for PCPs on enhancing communication of clinical empathy and realistic optimism.(14, 18, 19) Twenty-five primary care practices were randomised to access the EMPathicO e-learning package (115 PCPs) and 28 to consulting as usual (118 PCPs). Before signing up for the trial, patients booked consultations (for our purposes, an “index” consultation) as usual with a PCP (e.g. GP, FCP, Nurse Practitioner) and were unaware of whether their practice or PCP was randomised to have access to EMPathicO. Adults ≥18 years with an appointment to consult a participating PCP in-person, by telephone, or videoconference were then recruited into two groups prior to consultation: those consulting about new, recurrent, or ongoing MSK pain and scoring ≥ 4 on an 11-point scale at baseline;(20) and those consulting for any other reason (All-comers). Patients completing online consults were not eligible to take part in the trial. Eligible participants were invited to complete consent and baseline measures online prior to their “index” consultation; at this point, participants were invited to also give consent to contact for an interview to explore their views and experiences of their index consultation.(14)

We invited 120 trial participants to take part in an interview and emailed them a participant information sheet describing the interview study. The 120 invitees were purposively sampled using self-reported demographic data from 697 EMPathicO trial participants consenting to be contacted about taking part in an interview. We used maximum variation sampling (21) to capture diversity along dimensions of characteristics including age, gender, ethnic background, employment status, level of education, and index of multiple deprivation (IMD) score. We also sampled by reason for consulting (MSK pain or other conditions), index consultation method (in-person or telephone), PCP (GP, Physiotherapist, Nurse Practitioner), and primary care practice. The researcher telephoned participants to build rapport, answer questions, and arrange a mutually convenient date for the interview within 7-14 days of their index consultation. At the time of interview, some participants may have completed follow-up questionnaires as part of the trial (see Bishop et al., 2025(18)).

### Data collection

Interviews were conducted by RDH, SP, and ET; and ID and SF (female students completing an MSc in Health Psychology). Interviewers were blinded to RCT allocation group. A semi-structured topic guide [see Additional File 1] comprised open-ended questions to explore participants’ experiences of their index consultation, including expectations and outcomes from the appointment, as well as aiming to identify any perceptions of empathy and optimism during that encounter. We also explored the wider context of consultations in primary care, including views of arranging appointments and consultation modality. Interviews were audio recorded using MS Teams and transcribed verbatim. Interviews typically lasted 30 minutes (range 12-55 minutes). At the end of the interview, participants were debriefed, offered a copy of the interview transcript, and given a £10 voucher to thank them for their contribution. Interviewers completed field notes at the end of each interview. Thirty-nine interviews were conducted between March – June 2023. Data collection was then paused to allow for familiarisation with the data and identification and development of a thematic framework. It also enabled sampling from a wider range of practices. A further 32 participants were interviewed between August – December 2023.

### Data analysis

Data analysis occurred concomitantly to data collection, using the Framework Method.(16, 17) RDH led the analysis, familiarising herself with the transcripts. The transcripts were both deductively and inductively coded. Deductive coding included behaviours indicative of clinical empathy and realistic optimism that were delivered to PCPs as part of the EMPathicO training,(5) along with any *a priori* issues that were informed by the original research aims (to identify patients’ perceptions of empathy and optimism in primary care consultations) and questions from the topic guide (such as issues around access to primary care consultations, continuity of care, and perceptions of health after the consultation). Finally, inductive coding was applied to other relevant data and was supported by regular meetings with the qualitative team to discuss observations and reflections. A working analytical framework was developed [see Additional File 2] and applied by indexing data from subsequent transcripts. NVivo12 (22) was used to facilitate the coding, indexing, and charting of data to the framework matrix. The charted data were interpreted and developed into themes. Qualitative comparisons were made between participants in the MSK and All Comers groups and trial arms, but no clear or distinguishing patterns were identified from the data.

## Results

Seventy-one participants agreed to be interviewed (others did not respond to the invitation (n= 43), were unavailable due to work or family commitments (n= 4), had their index consultation cancelled (n=1), or were too unwell (n=1). A summary of participant characteristics is shown in Table 1.

**Table 1.** Participant Characteristics.

| Participant Characteristic | N (%) |
| --- | --- |
| **Age** |  |
| Range, Median | 24 – 85, 63.0 |
| **Gender** |  |
| Female | 44 (62.0%) |
| Male | 27 (38.0%) |
| **Ethnicity** |  |
| White British/Irish | 62 (87.3%) |
| African | 1 (1.4%) |
| Caribbean | 1 (1.4%) |
| Indian | 4 (5.6%) |
| Multiple Ethnic Groups | 2 (2.8%) |
| Persian | 1 (1.4%) |
| **Employment** |  |
| Employed | 31 (43.7%) |
| Retired | 36 (50.7%) |
| Unemployed/Unable to work | 4 (5.6%) |
| **Education** |  |
| University | 41 (57.7%) |
| School & College | 26 (36.6%) |
| No formal qualification | 4 (5.6%) |
| **IMD Score** |  |
| Range, Median | 2 – 10, 6.5 |
| **Reasons for consulting** |  |
| MSK pain | 32 (45.1%) |
| Other conditions | 39 (54.9%) |
| **Index consultation method** |  |
| In-person | 51 (71.8%) |
| Telephone | 20 (28.2%) |
| **PCP** |  |
| GP | 53 (74.6%) |
| First Contact Practitioner | 12 (16.9%) |
| First Contact Practitioner & GP | 3 (4.2%) |
| Nurse Practitioner | 2 (2.8%) |
| Nurse Practitioner & GP | 1 (1.4%) |
| **Practice** |  |
| Intervention Arm (16 practices) | 35 |
| Control Arm (13 practices) | 36 |

Most participants talked positively about their consultations and described interactions where they felt listened to, understood, and cared for. However, we also identified that these perceptions were impacted by broader contextual issues. Four themes captured the essence of participants’ experiences and perceptions of empathy and optimism in primary care: a stretched and inefficient primary care system; being known: PCP preparedness as a marker of empathy; being seen: consultation modality shaped perceptions of empathy; and knowing the next steps: realistic optimism through action. Shared across themes were concepts of time relating to patient effort, perceived pressure on PCPs, and waiting for onward referral and care, and its influence on perceptions of empathy and optimism.

### A stretched and inefficient primary care system impacting empathy and optimism

Experiences of appointment booking systems could influence how participants perceived empathy within primary care systems. Participants acknowledged that completing online consultation forms (such as eConsult) prior to an appointment could provide opportunities to describe concerns in their own words and enabled autonomy in deciding which PCP they would like to see, facilitating a sense of continuity of care. Others, however, commented on how standardised questions would not always fit with their specific symptoms or reasons for consulting. Participants felt limited in being able to fully explain concerns in their own words, which could make them feel less optimistic that they would be directed towards appropriate care. Having to communicate in writing without opportunities to speak to someone meant there were further barriers to concerns being fully understood, as well as limiting choice as to which PCP participants wanted to see:

> *I don’t particularly want to go and see [the physiotherapist] […] so although you’re asking to see the doctor, the nurse or whoever will triage it, will see it’s knee pain or back pain, and put you in to the physiotherapist […] I wanted to see my doctor. (73115, Physiotherapist, In-person)*

Participants also alluded to the time and effort it would take to complete the online consultation form, for the system to then categorise their request for care according to ‘best fit’, rather than paying attention to their specific concerns and expectations:

> *I first went into the category along the lines of managing a skin condition. Then it told me to tell them about the rash, which I did not have. It’s completely ridiculous. So I had to go through another category which was ‘change to medication’, which is not exactly this, but it was the closest one. (75939, GP, In-person)*

With regards to using telephone booking systems, participants commented on the frustration of having to wait in a queue before speaking with a receptionist. After investing time to get to the front of the queue, participants then felt a perceived lack of control or autonomy around how their disclosure of personal problems would be managed by the receptionist. Disclosing private concerns to administrative staff could feel disempowering and undermined the clinical empathy they experienced in consultations when relaying the same information; but participants felt it was necessary as a means to secure a consultation:

> *Back in the day, I would have rankled at telling the receptionist my own personal medical problem […] in all honesty I wanted not to waste reception’s time, or my time. Yes, I was up front about what the issue was and then I got an appointment clocked in. (34250, GP, In-person)*

Participants also reflected on the length of time it took between booking the appointment and having it. Despite system pressures leading to longer waits, many talked about the value of ongoing interpersonal continuity of care, and would opt to wait longer to see a PCP they had previously seen:

> *She’s my usual GP and I really like her, so I try whenever possible… I do have an option when I phone the receptionist to speak to a different GP, who is more available, but I choose to wait for my own. (34949, GP, Telephone)*

In contrast, other participants felt that continuity of care was not possible due to the need for primary care systems to meet demands for consultations, so securing any appointment was better than nothing:

> *In my surgery, you can’t really choose who you see, so I was just happy to have an appointment with a GP to be honest! [chuckles] (24169, GP, In-person)*

Overall, participants’ accounts suggested that the process of getting an appointment could shape perceptions of empathy before the consultation began. The *“need to go through certain hoops”* (*27289*) to get an appointment left some patients frustrated, feeling less treated as individuals, and with limited autonomy over their care preferences, even when the subsequent consultation with the PCP was experienced positively.

### Being known: PCP preparedness as a marker of empathy

Participants’ perceptions of PCP ‘preparedness’ influenced their experiences of empathy during consultations. Having, and clearly communicating, some knowledge of a participant’s personal background, medical history, or reason for consulting was experienced favourably:

> *She’d obviously read my notes before I went in. […] That made me feel that she was actually thinking about me as an individual rather than just another patient. I appreciate that really, because obviously they’ve got a lot of patients to see […] It was good that I felt she knew what it was that she needed to do. (20021, GP, In-person)*

This preparedness was perceived by participants as a marker of empathy, even if it resulted in being called later than their scheduled appointment time. Not reading any notes beforehand could set a negative tone, particularly when PCPs were behind schedule. A lack of preparedness implied a perceived lack of interest from the PCP, and limited opportunities for empathic communication within a time-limited consultation:

> *He was already running 10 minutes late […] And I know they see hundreds or thousands of people a week or month, but I just felt like he should have had a quick look at my notes from my last appointment and then thought, oh, actually, yeah. I asked her to book in. Just not asked me what he could do for me. (82858, GP, In-person)*

Participants also highlighted how the opening of the consultation could shape their sense of being known and cared for. A personal greeting, such as using the participant’s name, could work to put them at ease and make them feel that there would be time and space for discussing concerns and potential treatment during the consultation:

> *He actually greets you, says ‘Hello,’ calls you by your name. It just makes you feel easy enough to talk to him. (42488, GP, Telephone)*

These behaviours were described as directly helping consultations feel less like *“a case of getting me in the appointment, and getting me out to get the next patient in.”* (*26621*)

### Being seen: consultation modality shaped perceptions of empathy and optimism

While participants were generally positive about their consultation experiences, context around the type of appointment (i.e. over the telephone or in-person) could influence perceptions around feeling cared for.

Participants seemed more understanding and accepting towards PCPs who were running behind schedule for in-person appointments; as being physically present in a primary care setting provided a sense of control and reassurance that they would have an appointment. On the other hand, uncertainty around knowing when a PCP would call for a telephone consultation could create worry about missing the call, or whether they would have the appointment at all:

> *They tell you, ‘oh, the doctor will ring you between 12:00 and 3:00.’ And you’re waiting for the phone call for like 3 hours. Whereas when you’re face to face, you’ve got an appointment at 12:00, then he might be running 20 minutes late, but you’re there and you know that you’re going to be seen. (82858, GP, In-person)*

These experiences of waiting for telephone calls without a definitive appointment time led some participants to feel that the system was not empathic towards their individual circumstances, as they were expected to remain available at any time, potentially disrupting work, social, or caring responsibilities.

Telephone consultations were perceived as *“better than nothing” (20021, GP, In-Person)* given the difficulties in getting appointments. Some participants felt the care they received was less empathic over the telephone, and had “*not been lasting interactions, [but] one-off consultancies” (65763, GP, Telephone).* PCPs appeared busier or more pressured to keep to time during telephone appointments, leading some to worry about becoming a burden to both the PCP and the primary care system:

> *I’m always conscious of time over the phone […] I sort of think ‘oh gosh I’m keeping somebody waiting’ if I go over. I think we’re allowed ten minutes over the phone, and I’m very conscious of that because I don’t like to keep the next one waiting. (76855, GP, In-Person)*

In this context, having an in-person appointment could feel like a ‘step-up’ from a telephone consultation for some participants, fostering a greater sense of optimism and ongoing care:

> *I felt lucky, just being able to see a doctor, and not talking over the phone, that’s great […] I actually thought, I’m getting somewhere. (27289, GP, In-person)*

### Knowing the next steps: realistic optimism through action

Participants’ perceptions of optimism were strengthened when PCPs not only clarified what action would be taken, but how this action sat within the broader pathway of care. This included discussion and shared-decision making around treatment options, arranging tests or referrals, agreeing to follow-up with participants, or safety-netting if participants wanted further support.

Shared decision-making around care and taking action during a consultation fostered a sense that the PCP understood the participant’s concerns and was working with them to improve symptoms or quality of life in a timely manner:

> *[The GP said] ‘I think I’ll have a specialist look at it, as long as you agree?’ I said I do agree. That’s what I want to hear […] She started typing up the referral and told us that she was going to send it off straight away when the appointment had finished. (13833, GP, In-person)*

Participants felt optimistic when they had a structured plan for their treatment, particularly when they would need to be engaged with primary care services over a period of time:

> *I think the fact that she outlined a series of steps that were going to be taken was particularly helpful […] It’s clear to me that it’s not something that’s going to be cleared up in just one appointment. It’s going to take a couple of months to get to the bottom of this, but at least there is a plan of action. (99283, GP, In-person)*

Honesty was valued by participants if PCPs were uncertain around the diagnosis, treatment, or outcome for their concern. Trust and perceptions of care could be maintained if PCPs were able to explain their decision-making and outline a plan to support the participant moving forward:

> *She clearly said what she knew, and what she didn’t, and where the width of her knowledge stopped […] and that was very honest […] she said, ‘Well, there’s an expert [at another clinic] and she’s asked me to refer anyone who might have specific issues.’ […] So yes, I think it was approached in a very open way in the conversation, and it was my decision to make that call or not. (75939, GP, In-person)*

Conversely, participants found it harder to feel optimistic about their care when they were unsure about what would happen next, or if a course of action had been recommended but not explained. One participant presenting with MSK pain was confused after their encounter with an FCP:

> *Weirdly, at the end of the appointment, she said, ‘Oh, I’m going to refer you to a physio’, and I should have asked at the time, but I was like, ‘I thought you were the physiotherapist.’ She absolutely was because I’m pretty sure that’s what it said on her room, and then she knew what she did. She talked like a physiotherapist, so I should have said, ‘Okay, what’s the difference between this next appointment I’m going to have and you?’ (42175, Physiotherapist, In-person)*

Participants felt reassured when PCPs were clear that they could return if symptoms persisted, their treatment was not working, or they had any additional concerns. This helped participants feel that their care could continue over time, rather than ending at the close of the consultation:

> *She’d opened the door that if I’m feeling dissatisfied, I can ring up and book an appointment […] I felt reassured that there was a follow-up. I wasn’t just left to – ‘You’ve got what you want, off you go’. It’s a process - she actually used the word, it’s a ‘journey’. (29708, GP, In-person)*

However, a need to re-start the process of booking an appointment with a PCP and having to navigate the primary care system could undermine the sense that they would be able to easily access ongoing care:

> *In retrospect I should have asked for a follow-up appointment, because once again, that is the really difficult part. (99283, GP, In-person)*

Participants also recognised that, in a stretched system, PCPs could not always provide immediate resolutions, but being told what would happen with results, when follow-up might occur, or what options would be discussed next could still feel reassuring. However, inefficiencies with communication within the primary care system could undermine the actions PCPs took to elicit perceptions of empathy and optimism during the consultation:

> *He said I’d receive a text message when I’d been referred [to a specialist] […] I still haven’t had a text message, and knowing, unfortunately, the state of the NHS, I think once I’m referred it could be months before I see somebody. (73984, MSK Pain, GP, In-person)*

## Discussion

This study shows that patients’ perceptions of empathy and optimism are not only shaped by PCPs during the consultation, but also by the primary care system within which they work. Patients acknowledged that PCPs were “doing their best” within a stretched system, but unless wider organisational systems support empathic care, these pressures may reduce PCPs’ willingness or capacity to be empathic, as well as limiting patients’ opportunities to perceive, and benefit from, empathic care.(4) There is a need to focus on fostering and maintaining *empathic systems*; where systems and institutions are structured in a way that facilitate empathic interactions throughout the whole health service.(23) Participants’ accounts highlighted three mechanisms through which this occurs: accessing care, the modality of the consultation, and the organisation of care beyond the consultation.

Patients want access that is timely, responsible, understandable, and appropriate to their needs.(13) While evidence suggests more digitised access systems (such as telephone triage and online consultation platforms) help to meet demand and are convenient and efficient for patients (24, 25), our findings emphasise the need for “human fit” within these systems if there is to be further digitisation of the NHS system.(26) As reflected in our study and other research, if patients spend a disproportionate amount of time and effort negotiating ‘loops’, ‘hurdles’ (27), and ‘hoops’ when accessing appointments, yet are still unable to explain individual concerns, choose continuity of care, or understand triage decisions, perceptions of empathy and realistic optimism from the PCP during consultations could be diminished.

The structure and modality of consultations may also shape perceptions of empathy and optimism. While empathy can be communicated effectively in remote consultations (28, 29) our findings imply that some interactions over the telephone may feel more transactional and time-pressured (30), limiting opportunities for patients to ask questions or raise concerns, and leading some to feel they are creating additional burden on an already-stretched system. If primary care systems are to use both in-person and remote approaches to meet patient demand for appointments, it is important they can be adopted in a timely and efficient way, to ensure that PCPs’ time during consultations facilitates the delivery of empathic care, rather than delivering care that is fragmented, transactional and less patient-centred.(8)

Patients’ perceptions and experiences of empathy and optimism go beyond the consultation and are influenced by how care pathways are communicated and organised over time. Participants in our study described feeling optimistic when PCPs provided clear treatment plans, transparently explained and accounted for uncertainty, and outlined next steps in their care, where appropriate. This emphasises that perceptions of empathy are temporal and highlight the importance of organisational, interpersonal, and informational continuity of care, both within primary care and if patients require referral to secondary care. Our findings show that timely action from PCPs during the consultation such as referral and safety-netting advice can enhance perceptions of empathy. However, these perceptions can be modified by system issues, such as a lack of communication or longer waiting times to be seen in secondary care, which has been previously reported in the literature.(31, 32)

System-level factors such as fostering a culture of empathy, sensible workloads, an appropriate physical environment and adequate time can make a difference to the delivery of empathy in healthcare (33), therefore organisations need to be structured to facilitate PCPs’ capacity to communicate empathically during consultations.(23, 32, 34) However, access systems appear to be designed at a practice-level and are less patient-centred (35), and increasing pressures on PCP workload could lead to burnout, which in turn could lead to a decline in empathic practice during consultations.(32) The NHS 10-year plan for England (36) acknowledged a need to “provide continuous, accessible and integrated care” from hospital to community settings. The plan hopes to “revitalise access to general practice” by utilising the NHS App to act as a “doctor in their pocket”; allowing patients more autonomy to book appointments and communicate with PCPs, as well as access advice, review care plans, and self-refer to local tests and services. However, in line with the existing evidence (8, 23, 26) our research further emphasises the need to address system-level issues that promote or hinder empathic practice. Training at an early stage (i.e. during undergraduate medical studies) for a range of healthcare practitioners, and training and support for administrative staff could be one approach to equip the primary care workforce with sustainable communication approaches that meet a stretched, pressured, and evolving healthcare landscape. Otherwise, there is a risk that further operationalisation of care provision (such as the use of Artificial Intelligence), along with the need to meet targets and protocols, could threaten clinical empathy and realistic optimism in primary care consultations.

### Strengths and Limitations

This study was nested within a large, multi-centre, cluster-randomised controlled trial, which allowed for exploration of patient experiences across a diverse range of practices and consultation types in primary care settings. The use of maximum variation sampling and a large sample size allowed for in-depth exploration of a wider range of perspectives. Additionally, blinding of both participants and interviewers to trial allocation helped ensure that accounts reflected routine experiences of care. Participants were invited to discuss empathy and optimism in general as well as reflect on specific aspects of their index consultation, and we are confident in the relevance, quality and importance of the data gathered.

Participants were predominantly from a white ethnic background and had higher levels of education, which may limit the transferability of findings to populations more likely to experience health inequities, including those facing language barriers, lower health literacy, or digital exclusion. Participants were sampled from primary care practices in England and Wales, meaning the experiences may not necessarily be reflective of primary care systems in other countries.

## Conclusions

Patients’ perceptions of clinical empathy and realistic optimism are not only shaped by PCPs’ communication during consultations, but also by the systems through which care is accessed and delivered. Challenges and pressures around accessing appointments, communication according to consultation modality and navigating uncertainty after consultations can undermine patients’ perceptions and experiences of empathy in consultations. Organisational structures and workforce development should therefore treat empathic communication as both an individual skill and system-level responsibility, ensuring that organisational processes facilitate, rather than impede, positive patient-practice relationships within primary care.

## Data Availability

Data are available on request. Requests for deidentified participant data may be submitted to the University of Southampton data repository, quoting doi: [to be added on acceptance]. Requests would be subject to review by a subgroup of the trial team. Access to anonymised data may be granted following this review, subject to conditions including ethical approval, qualifications, and aims consistent with the original purpose of the study. All data-sharing activities would require a data-sharing agreement.

## Abbreviations

TIP: Talking in Primary Care
PCP: Primary Care Practitioner
FCP: First Contact Practitioner
IMD: Index of multiple deprivation

## Additional Files

File Name: Additional File 1

File Format: .pdf

Title: TIP Patient Interview Guide

File Name: Additional File 2 File

Format: .pdf

Title: TIP Analytical Framework

## Declarations

### Ethics approval and consent to participate

The study was conducted in accordance with the Declaration of Helsinki. Ethical approval was given by the South Central Hampshire B Research Ethics Committee (1^st^ July 2022) and the Health Research Authority and Health and Care Research Wales (6^th^ July 2022) (REC: 22/SC/0145; IRAS ID: 312208). Informed consent was audio recorded and electronically signed prior to interviews.

### Consent for publication

Participants provided informed consent for the publication of anonymised quotations.

## Funding

This project was funded by the National Institute for Health Research (NIHR) School for Primary Care Research grant (project reference 563). The Primary Care Research Centre, University of Southampton is a member of the NIHR School for Primary Care Research and supported by NIHR Research funds. Service support costs were paid by the CRN. CDM is funded by the National Institute for Health Research (NIHR) Collaborations for Leadership in Applied Health Research and Care West Midlands and the NIHR School for Primary Care Research. The EMPathicO e-learning package was developed using LifeGuide software, which was partly funded by the National Institute for Health Research Southampton Biomedical Research Centre (BRC). NIHR Local Clinical Research Networks (CRNs) supported practice recruitment.

The views expressed are those of the authors and not necessarily those of the NIHR or the Department of Health and Social Care.

The study sponsor (University of Southampton) and funders had no role in study design; collection, management, analysis, and interpretation of data; writing of the report; or the decision to submit the report for publication. The researchers are independent from the funders and all authors had full access to all of the data (including statistical reports and tables) in the study and can take responsibility for the integrity of the data and the accuracy of the data analysis.

### Authors’ contributions

All authors meet the ICMJE criteria for authorship, i.e., have made substantial contributions to the conception or design of the work; or the acquisition, analysis, or interpretation of data for the work; AND drafted the work or reviewed it critically for important intellectual content; AND approved the version to be published; AND agree to be accountable for all aspects of the work in ensuring that questions related to the accuracy or integrity of any part of the work are appropriately investigated and resolved.

Specific contributions as per the CRediT taxonomy are: Conceptualization (FLB, TB, KG, NC, RDH, ET, AH, MER, MJR, CDM, JB, BS, LM, SP, JV, HA, JH, GML, JN, NI, PHL, PL, HAE); Data curation (RDH, SP, ET, ID, SF); Formal analysis (RDH, ET, FLB, GML, LM, SP, HAE); Funding acquisition (FLB, KG, MJR, CDM, JB, BS, LM, JV, HA, JH, GML, PL, HAE), Investigation (NC, RDH, ET, AH, MER, SP, NI), Methodology (FLB, ET, HAE), Project administration (FLB, NC, HAE, JN), Supervision (FLB, MJR, CDM, JN, HAE), Writing – original draft (RDH), Writing – review & editing (ET, SP, GML, HAE, LM, HA, JH, ID, SF, JB, JV, NC, PL, CDM, MJR, AH, MER, JN, TB, KG, BS, NI, PHL, FLB).

The guarantor (F L Bishop) accepts full responsibility for the work and/or the conduct of the study, had access to the data, and controlled the decision to publish. The corresponding author attests that all listed authors meet authorship criteria and that no others meeting the criteria have been omitted.

## Acknowledgements

The TIP trial team gratefully acknowledge the contribution of our Public Advisory Group (Jennifer Bostock, Clara Martins de Barros, Mark Lamond, Manoj Mistry, Hazel Patel, and David Truswell), our Independent Trial Steering Committee (Joanne Reeve, Ian Dickerson, Ines Rombach, Philip Pallman), and our administrator (Tanya Palmer).

