## Additional file 1 for "System pressures may threaten patient perceptions and experiences of empathy in primary care consultations: A nested qualitative interview study"

### Patient Interview Topic Guide
### Initial Consultation

***Iteration and Flexibility***

*The topic guide will evolve as the interviews progress in response to researchers' reflections, initial analysis, and feedback from participants. The scope of the interviews will always remain consistent with the objectives specified below. Within this scope, the topic guide will be used flexibly in that the researcher may adjust the questions asked and the order in which they are asked as appropriate for the individual participant. It may not always be necessary to ask all of the questions in order to elicit the relevant information.*

***Objectives***

*1. To explore patients' views, experiences and insights of their initial consultation and subsequent consultations with primary care practitioners taking part in the TIP Trial.*

*2. To explore patient's perceptions of the initial consultation and its subsequent impact on their health.*

*3. To explore patients' experiences of taking part in the TIP Trial.*

#### Topic Guide

*Clarify with participant if they saw a GP, nurse, or physiotherapist. For the purpose of the topic guide, "PCP" will be used but should be changed to the relevant healthcare provider during interviews.*

**a.** Could you tell me all about your recent appointment with your PCP?

*That's really interesting. I'm now going to ask you a little bit more about some of the things you've just talked about.*

**b.** What led you to make this appointment?

- What were you hoping for from your appointment?
  (Prompt: was patient seeking diagnosis, examination, advice, treatment, referral, prescription?) Did you have any ideas of what the problem was/what you were expecting?

**c.** What happened during the appointment? (**If they mention specific clinician behaviours – follow-up with - and how did that make you feel?**)

- How did you feel about the way the PCP talked to you during your appointment?
- How did you feel about the way the PCP listened to you during your appointment?
- How did they answer any questions you had?
- How did they respond to any concerns you had?
- Could you describe anything the PCP said or did that you found to be helpful?
- Could you describe anything the PCP said or did that you found to be unhelpful?

- How did the decision to [refer/prescribe/other outcome] come about in the consultation? Were you involved in that decision making process? (How?)
- What do you think about the diagnosis/advice/treatment/information/referral/prescription the PCP gave you?
- What's your understanding of why you have come away with this diagnosis/advice/treatment/information/referral/prescription?
- What do you think the diagnosis/advice/treatment/information/referral/prescription might do for you? (Prompt: any changes in symptoms, wellbeing, coping)

Did the PCP do or say anything in particular that made you feel cared for? (Probe what they did)

Did the PCP do or say anything in particular that made you feel hopeful? (Probe what they did)

*(If applicable)* Could you tell me about having an interpreter in the appointment with you?
- Who was in the appointment with you, was it a family member, or someone else?
- How did the doctor talk with you about your problem? Did they talk directly to the interpreter or you?

**d.** Can you tell me about the PCP you saw this time?
- Is this someone you've seen before?
- How did you feel seeing this PCP?

**e.** Could you remind me how you had your appointment (face-to-face, telephone, video)?
- How did you book your appointment (eConsult, reception)?
- How did you find booking the appointment?
- How did you feel about having your appointment this way?

**f.** Overall, how did you feel after your appointment with the PCP?
- What did you get out of your appointment?
- Do you feel you had enough time for your appointment? Why/Why not?
- How would you feel about seeing this PCP again?
- How do you feel about your health now, compared to before?

Wrap up question:

**g.** Is there anything else you'd like to tell us about your appointment that we haven't talked about, that you think may be important for us to know?

Before we finish for today, I'd like to ask you a few questions about taking part in this study.

What encouraged you to take part in this study? (Explore positive and negative views about this study – Did you think there would be any benefits? Did you have any concerns or doubts about taking part?

How have you found filling in the questionnaires so far? If there anything we could do to improve that part of the study?

Is there anything else you would like to share about your experiences of taking part in this study?
