## Additional file 2 for "System pressures may threaten patient perceptions and experiences of empathy in primary care consultations: A nested qualitative interview study"

**TIP Qualitative Interviews (Patient) Framework**

*Framework has been developed and created deductively, including behaviours indicative of clinical empathy and realistic optimism that were delivered to PCPs as part of the EMPathicO training (marked with a '#'); any a priori issues that were informed by the original research aims to identify patients' perceptions of empathy and optimism in primary care consultations; and questions from the topic guide (such as issues around access to primary care consultations, continuity of care, and perceptions of health after the consultation)*

| Name | Description |
|---|---|
| 1. Reason & expectations for appointment | |
| 1.1 Reason for appointment | Participant talks about the reason for having an appointment |
| 1.2 Patient history | Patient talks about history leading to making an appointment |
| 1.3 Expectation from appointment | Participant was talking about what they were hoping for from their appointment |
| 2. What happened during appointment | |
| 2.1 Second concern | Participant mentions a second concern raised at appointment |
| 2.2 Partner or second person in appointment | Participant describes the presence of another person in the appointment |
| 2.3 Only treating one problem area | Participant talks about how they are only treating one problem area, not a holistic approach |
| 3. Perceptions of Empathy | |
| 3.1 # Alleviating concern | The explanation of treatment outcome alleviated patient concern |
| 3.2 # Body orientation and gaze | PCP faces and looks at Participant |
| 3.3 # Computer use | Participant talks about computer use during the consultation |
| 3.4 # How condition affects patient's life | PCP asks Participant how condition is affecting their life |
| 3.5 Ask Participant about symptoms | PCP asks Participant about the symptoms they are experiencing |
| 3.6 Treating participant as an individual | Participant talks about PCP treating them as an individual |
| 3.7 # PCP checking understanding | Participant talks about whether the PCP has checked any understanding of what's been said |
| 3.8 # Affirmation that concerns are valid | PCP shows behaviour that makes Participant feel that their concerns are valid |
| 3.9 # Positive body language | PCP uses positive body language |
| 3.10 # Referring to goals during the consultation | PCP refers to Participant's goals during the consultation |
| 3.11 Feeling cared for | What made the Participant feel cared for |
| 3.12 How PCP listened | Participant talks about how the PCP listened to them |
| 3.13 How PCP talked | Participant talks about how the PCP talked to them |
| 3.14 Asking questions | PCP asks the Participant what they are currently doing to manage their condition |
| 3.15 Following up with patient | How the PCP followed-up with them with advice/treatment after index consultation |
| 3.16 Small talk or humour | Participant talks about making small talk or use of humour during the consultation |
| 3.17 PCP shares their own experience | PCP talks about their own experience to offer a different perspective on Participant's condition |
| 3.18 Taking Participant seriously | Participant talks about how they felt they were being taken seriously by the PCP |
| 4 Perceptions of Optimism | |
| 4.1 # Explanation of the treatment and how it works | PCP gives an explanation of the treatment and how it works |
| 4.2 # Clear instructions for the treatment | PCP explains how the procedure will be carried out |

| | |
|---|---|
| 4.3 Explaining benefits of treatment | Participant talks about any benefits of treatment described by the PCP |
| 4.4 Explaining risks of treatment | PCP tells participant there are risks to treatment |
| 4.5 # Expected outcomes of treatment | What PCP tells Participant what they can expect from treatment |
| 4.6 Participant tells PCP what needs to be done | Participant tells the PCP what they are expecting based on previous consultations and/or advice |
| 4.7 # Positive words to describe treatment | PCP uses strong words to describe treatment |
| 4.8 # Relevant experience of clinician | PCP talks about their relevant experience, Participant feels that the PCP is knowledgeable |
| 4.9 GP is knowledgeable | Participant felt that the GP was knowledgeable |
| 4.10 Feeling hopeful | Anything the PCP said or did that made the Participant feel hopeful |
| 4.11 Explanation of results | Participant say that they felt reassured based on PCPs explanation of symptoms |
| 4.12 Empowering the patient | PCP gives participant treatment plan that gives them a sense of control over managing symptoms |
| 4.13 Hard to be optimistic because of condition | Participant talks about how it is hard to be optimistic when prognosis of their condition is poor |
| 4.14 No indication of treatment failure | PCP does not give any indication that treatment won't work |
| 4.15 Reassurance | How receiving reassurance makes the Participant feel reassured |
| 4.16 Receiving treatment | Receiving treatment makes the Participant feel optimistic that something is being done |
| 4.17 Seeking answers | Participant talks about still feeling unwell despite having tests and finding nothing wrong |
| 4.18 Unhelpful behaviour | Participant explains any behaviour from PCP that they found to be unhelpful |
| 4.19 No helpful advice | PCP does not give helpful treatment or advice to Participant |
| 4.20 Not feeling hopeful | Participants talks about not feeling hopeful |
| 4.21 PCP is not listening to Participant | Participant talks about how the PCP is not listening |
| 4.22 PCP disregards Participant concerns | The PCP has disregarded what the Participant has said to them and their concerns |
| 4.23 Participant feels dismissed | Participant feels dismissed by PCP |
| 5. Appointment outcome | |
| 5.1 # Safety netting | PCP gives Participant option to return if things get worse |
| 5.2 # Closing consultation positively | Participant talks about how the consultation was closed, considering verbal/non-verbal language |
| 5.3 Feeling after consultation | How Participant feels after their appointment |
| 5.4 Next steps | Participant talks about the next steps in their treatment |
| 5.5 Treatments not suggested by PCP | Participant is considering alternative treatments outside of what PCP has offered |
| 5.6 Decision making | Participant talks about how a decision was made for treatment/next steps |
| 6. Continuity of Care & PCP Characteristics | |
| 6.1 Seeing different PCP | Participant has been seeing different PCPs for the same health problem |
| 6.2 Seeing same PCP again | How Participant feels about seeing the same PCP again for future appointments |
| 6.3 Meeting PCP for first time | Participant talks about their experience of meeting a PCP for the first time |
| 6.4 Seen the same PCP before | Participant talks about how they have seen this PCP prior to the index consultation |
| 6.5 Age of GP | Participant talks about the age of their GP |
| 6.6 Gender of GP | Participant talks about the gender of their GP |
| 6.7 Participant shows concern towards PCP | Participant talks about how PCP was during appointment |
| 7. Access to care | |

| | |
|---|---|
| 7.1 Making primary care appointments | Participant talks about making appointments in primary care |
| 7.2 Type of appointment | Participant talks about how they had their appointment and views towards this |
| 7.3 Referral to secondary care | Participant has been referred to secondary care |
| 7.4 First contact practitioner | Participant offers views on seeing first contact practitioner within their practice |
| 7.5 Perceptions of primary care practice | Participants give overall views/Perceptions of primary care |
| 7.6 Reception giving test results | Participant talks about getting test results from receptionists |
| 8. Perceptions of health | |
| 8.1 Treatment has helped | Participant talks about how treatment/advice has been helpful |
| 8.2 Treatment has caused other problems | Participant talks about how treatment has caused other problems |
